# Physician-patient relationship at HUEH: is it a “relationship to” or a “relationship between”

**DOI:** 10.1101/2020.04.19.20065482

**Authors:** Axler Jean Paul, Yves Gardy Leonard, Rebecca Saint Louis, Jackyvens Camille, Hans Peter K. Delicat, Roubytherson Castra, Ricardi Jean, Rubens Delma, Yvelo Decan, Marc Felix Civil

## Abstract

To discover the relationship model in force between doctor and patient at the Haitian State University Hospital of Haïti (HUEH), a semi-directed survey was conducted among fifty patients. The qualitative analysis of the various interviews showed that patients were generally satisfied with their relationship with doctors. However, opinions are not sharing on their level information whether it is their illness or their therapeutic management; the results also showed that doctors had poor empathy. Hence our conclusion there is an unethical relationship in this institution, where doctors and patients coexist mainly in a “to” relationship and less so in a “between” relationship.

## Introduction

It is a fact that for a long time physician was perceived as the sole decision-maker in the management of patients, as evidenced by the Hippocratic oath, “I will direct the diet of the sick to their advantage according to my strength and judgment”, an oath repeated by health professionals of antiquity, which promoted the type of paternalistic relationship between physician and patient. Just as Platon, one of the leading philosophical and social thinkers of the time, had to say: let him be considered insane, any doctor who wants to explain his illness to a patient instead of just treating him [1]. The evolution of human and social sciences has brought to light the crucial importance of the patient’s participation in his or her care, and more particularly in decision-making. None can omit the words of Isaiah BERLIN, who said: I wish my life and my decisions to depend on me and not on external forces of any kind… I wish to be a subject and not an object [2].

The doctor-patient relationship, structured around demand and offer of care, a place of competence, method, rigor, and technique, is an intersubjective relationship, governed by affective springs which give it an extraordinary power of reciprocal influence and make it a major source of frustration and blockage, but also of mobilization and motivation for any long-term therapeutic project [3]. The evolution of the doctor-patient relationship can thus be thought of as a form of liberalization, which has become a topical issue. The doctor-patient relationship is of concern to us all, whether in the medical world (health professionals, students, etc.) or elsewhere; and this, in developed countries where public health affairs have enabled patients to group together in networks, associations, and even today to become civil parties in repeated lawsuits against hospital institutions [4], as in underdeveloped countries such as Haiti where, according to public opinion, the relationship between doctors and patients is unidirectional in health institutions and more particularly in the Hospital of the State University of Haiti (HUEH).

Considering the complaints, the lawsuits against doctors, health institutions and our lack of knowledge about the type of relationship existing at the HUEH, we conducted that research to find out whether it is a “to” or “between” type relationship. If the “to” relationship is thought on the model of an external perception that would be directly objective, the “between” relationship is thought on the model of feeling that progressively binds two beings [5]. The objective of this research is to contribute to an improvement in the quality of the relationship between doctors and patients through a better knowledge of the latter.

## Method

It is a cross-sectional, qualitative study with a phenomenological approach that was conducted over a period of eight months in Haiti’s largest health institution, HUEH. These patients are mostly of low socioeconomic levels such as shopkeepers, farmers, people unable to afford the care in clinics or other private facilities, or people referred to them for inadequate service from any other facility in the country offering care. Patients come from all age groups and different religions. In our research, patients were interviewed using a qualitative approach that promotes greater freedom of expression. Since the patient must be at the center of any management process, he or she is in the best position to assess the quality of his or her relationship with the doctor, for which they were the only ones maintained. To participate in the study patient had to be hospitalized or come to the hospital often, be in a conscious state, be aware of why he or she is being maintained and give his or her consent.

The data collection was done over a period of three days in all departments of the hospital except radiology, outpatient clinic, pathology, and ORL services. The data were collected using semi-directed interviews of six to ten minutes each, structured according to an interview grid evaluating different dimensions of the doctor-patient relationship from the time the patient was admitted to the department until the time of the interview, considering the patient’s level of knowledge of his or her illness, treatment, consideration for and expressions towards the doctor, and level of appreciation of that relationship. But also communication, decision making, availability, empathy and respect towards the patient.

We presented without our gowns to do not influence the patient’s statements. Apart from the grid, tape recorders were used to record the interviews with the patient’s consent, and software (word, excel) were used for the study. Poor quality records were eliminated and the remaining data were categorized according to semantic criteria and analyzed using the Colaizzi method [6]. The work was approved by the ethics committee of the Faculty of Medicine and Pharmacy of the State University of Haiti (LABMES: Laboratory of Ethical Medicine and Society).

### Results and Discussion

The results are presented in the form of tables grouping ten categories containing the most relevant units of meaning ranging from welcome to patient satisfaction.

Table 1 presents the characteristics of the patients in the study according to services, consent, gender and age groups. (Table.:1). If for “reception” category the frequency of appearance of the units of good *reception and* unsatisfactory reception *are* respectively 17 and 2, the categories “availability”, “respect”, “empathy”, “patient expressions”, and “patient considerations” have in majority units of meaning with the frequencies equal to the unit (Table.:2). For the “decision making” category, the frequencies of occurrence of the units vary from 2 to 3 (Table.:2). Whereas for the categories “communication” and “appreciation” the frequencies of appearance of the unit’s *satisfaction, confidence, information on the disease, uninformed patient, clearly understood* treatment, *good listening* is respectively 18, 15, 12, 8, 7, 6. (Table.:4)

**Table 1.** Population characteristics by service

| Services | Patients<br>approache<br>d | Patients<br>who<br>consented | Selected<br>Interview<br>s | F | M | Ratio | Age range |
| --- | --- | --- | --- | --- | --- | --- | --- |
| **Surgery** | 13 | 8 | 6 | 0 | 6 | 0 | Adults |
| **Dermatology** | 3 | 2 | 2 | 1 | 1 | 1 | Young and adults |
| **Maternity** | 7 | 6 | 4 | 4 | 0 | - | Young Ladies |
| **Internal Medicine** | 16 | 11 | 11 | 5 | 6 | 0.83 | Adults and the elderlies |
| **Ophthalmolog**<br>**y** | 6 | 5 | 5 | 2 | 3 | 0.67 | Youngs and adults |
| **Orthopedics** | 12 | 11 | 6 | 2 | 4 | 0.5 | Adults |
| **Pediatrics** | 8 | 5 | 4 | 3 | 1 | 3 | Children and Teenagers |
| **Urology** | 5 | 2 | 2 | 0 | 2 | 0 | Adults |
| **Total** | 70 | 50 | 40 | 17 | 23 | 0.74 |  |

**Table 2.** Categorization of patient reports on intake, management and decision making

| Reception |  | Supported |  | Decision-making |  |
| --- | --- | --- | --- | --- | --- |
| *Judgment* | *N* | *Judgment* | *N* | *Judgment* | *N* |
| Good reception | 17 | Correct | 3 | Without consent | 3 |
| Unsatisfactory | 2 | Attention | 3 | Consent sought | 2 |
| Greetings | 8 | Dedication | 1 | Decision not proposed | 3 |
|  |  | Negligence | 1 |  |  |

**Table 3.**
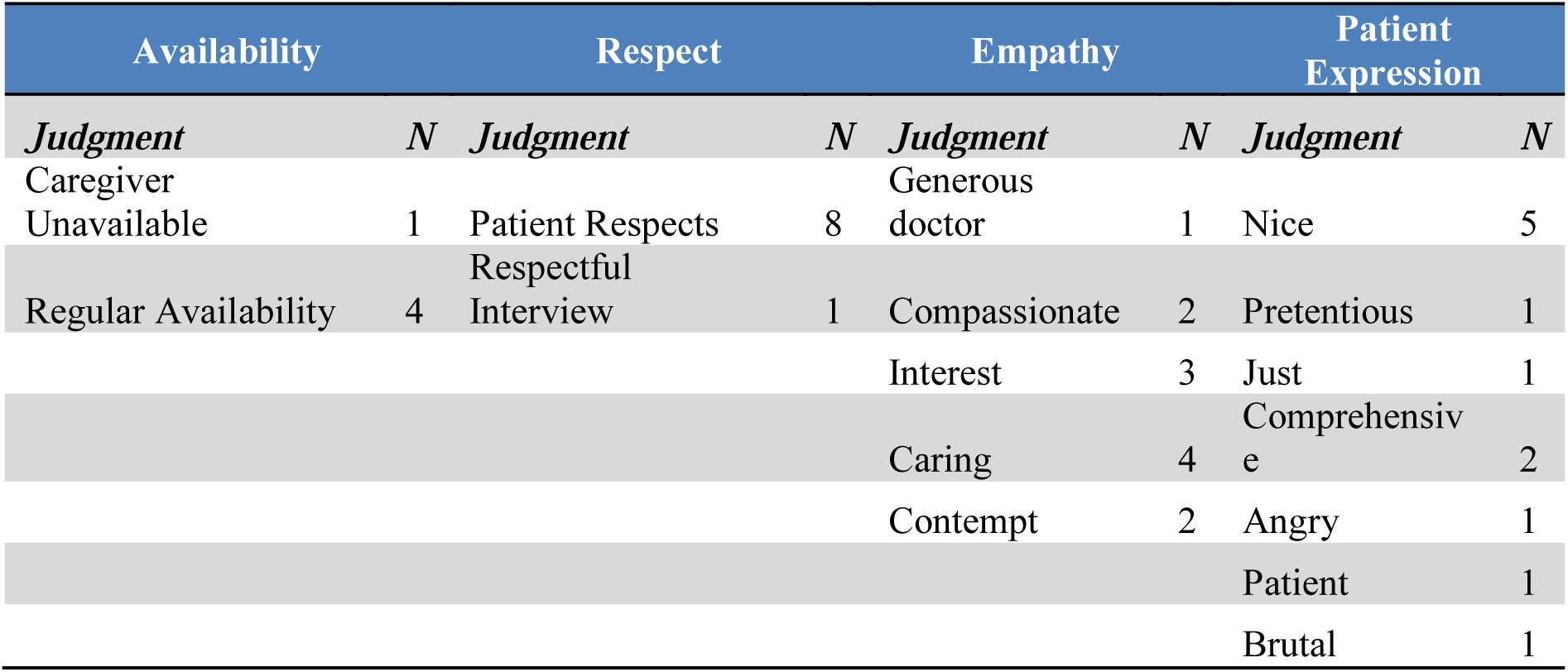
Categorization of patients’ statements about the physician’s availability, respect, and empathy, patient expressions

**Table 4.**
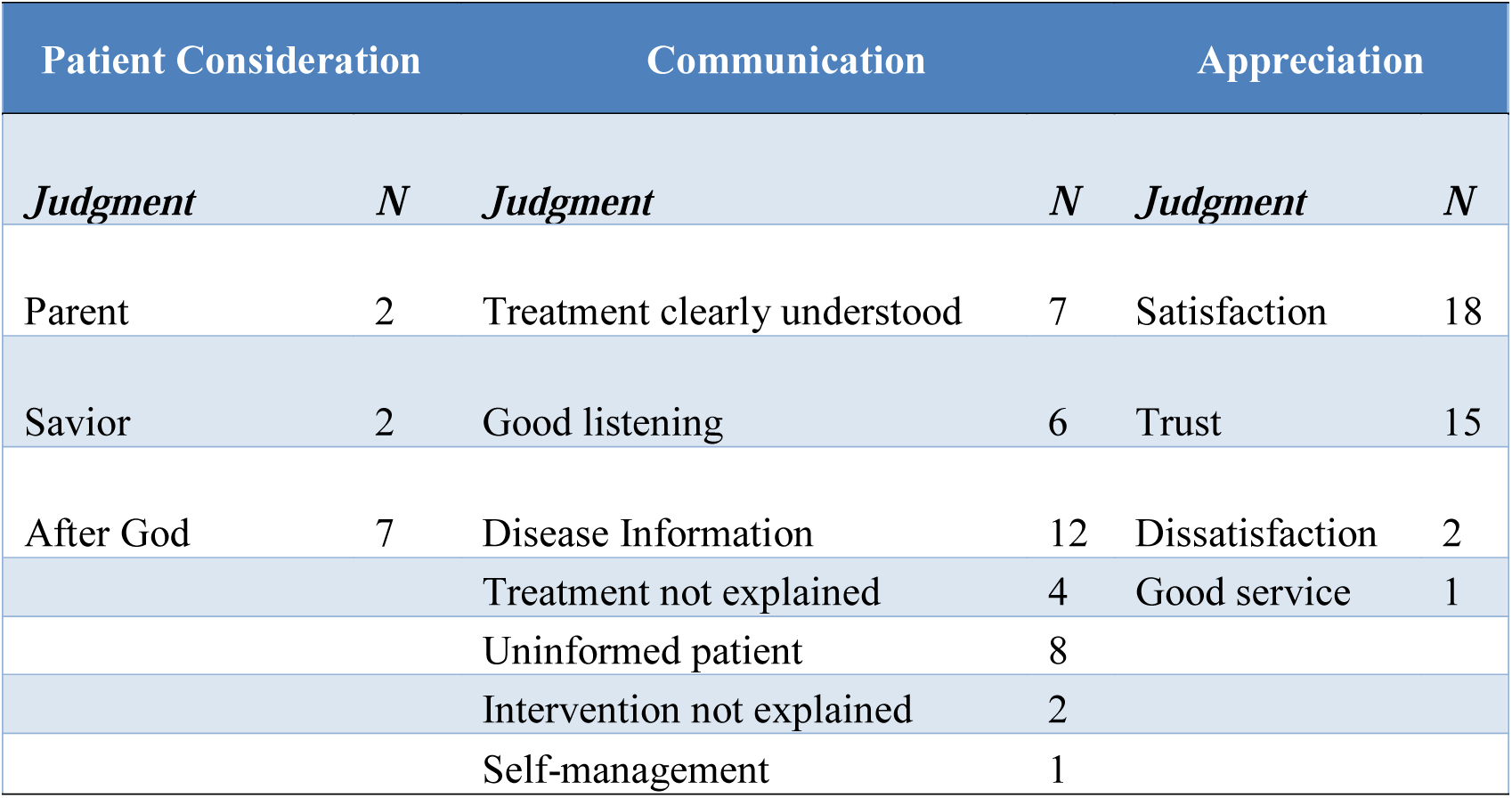
Categorization of patients’ statements about their consideration for the physician, their appreciation and communication between physician and patient

| Patient Consideration |  | Communication |  | Appreciation |  |
| --- | --- | --- | --- | --- | --- |
| *Judgment* | *N* | *Judgment* | *N* | *Judgment* | *N* |
| Parent | 2 | Treatment clearly understood | 7 | Satisfaction | 18 |
| Savior | 2 | Good listening | 6 | Trust | 15 |
| After God | 7 | Disease Information | 12 | Dissatisfaction | 2 |
|  |  | Treatment not explained | 4 | Good service | 1 |
|  |  | Uninformed patient | 8 |  |  |
|  |  | Intervention not explained | 2 |  |  |
|  |  | Self-management | 1 |  |  |

Reception plays an important part in the establishment of the doctor-patient relationship; it is above all a way of non-verbal but moreover verbal communication, meeting, and exchange. It conditions the smooth running of care, hence the interest to set the patient comfort as soon as he or she arrives [7]. There are three criteria to be considered when talking about quality care: *presentation, knowledge of the patient* and the guarantee of his or her *safety* [7]. On the other hand, we can see in the hospital, despite the low number of patients who say that the doctors present themselves to them at the time of reception, the majority are satisfied, with only two declaring themselves to be dissatisfied (Table.:2). The statements of these patients prove it:

> *“When I came to the hospital I was very well received, eight doctors took care of me”*. Patient of the Maternity ward.
>
> *“Sometimes I’m welcome, sometimes I’m not. Doctors don’t even introduce themselves to me, I hear their names mentioned by others.”* Patient of the Internal Medicine Dialysis Department

About the care, if the majority of patients declare it correct, we cannot, however, overlook the fact that few of them speak of neglect on the part of doctors, because a patient who feels neglected can become frustrated, which is no benefit to the relationship.

Some patients report that physicians seek their consent in making decisions. We know very well today the informed consent of patients is highly considered, based on the Kouchner law of 4 March 2002 in France and the Nuremberg Code was drawn up after the Second World War [8].

Thus, we observe the predominance of a paternalistic type of relationship between doctors and patients at the HUEH, a relationship which considers that in the name of the doctor’s mission, the patient’s consent is not medically relevant and should not be considered in this respect as a reference standard for the decision [9].

We could summarize the empathic attitude between physician and patient as the cognitive ability of the physician to put himself or herself in the patient’s situation while maintaining an emotional distance [10], although most patients generally prefer empathy to professional distance. Few patients report that physicians adopt empathic attitudes towards them, either by being generous or compassionate, such as this patient who said: “*When I was in pain, the doctor worried about me*. Orthopedic patient.

Others state the contrary, but it is not necessary to prove that the more empathetic the doctor is, the more the patient feels considered and the more he is ready to collaborate for better care of himself. As Miller found, nearly two-thirds of the differences in outcomes, in terms of six months of alcohol cessation, could be predicted by the degree of empathy shown by the workers during treatment [11].

According to the Public Health Code of France (Art L1110-2) [12], all health personnel must give the utmost consideration and attention to all persons, whatever their physical or mental state, culture, racial origin, political opinions and age [13]. In our study, few of them declare that they are respected, which calls into question the presence of respect between doctor and patient (Table.:3)

Moreover, whatever the state of mind of the doctor could be, it must not influence his relationship with the patient. It is the case of this patient who says about doctors: *Some are always in a good mood throughout the consultation but some others, perhaps disturbed or upset by personal problems, are not sometimes in a good mood*… *by my intelligence I have noticed a lack of balance*. Patient of the Dermatology department

At the Academy of Moral and Political Sciences, on January 30, 1950, Louis Portes declared: “in the exact meaning of the term the patient no longer sees clearly in himself, observing his evil and himself suffering from his evil, has slipped into opacity and sometimes even total darkness, all his steps in his knowledge of himself have become stumbling like those of a child” [14]. In this discourse of Portes, the image made of the patient helps us to understand the different perceptions of patients by their doctors, considering them in the majority of cases as envoys of God and sometimes as close relatives. Like those patients who say:

> *“Doctors are like men of God.”* Patient from Surgery
>
> *“For me there’s God and then the doctors and then my parents. After God, I trust the doctor*. “Patient of the Internal Medicine Department

Maintaining a relationship of exchange, this cannot be done only by revealing to doctor the different symptoms and giving the patient the necessary instructions to improve his health. Total information is one of the main issues in the doctor-patient relationship [15]. Information for patients and the care they receive is one of the central issues that run through the debates on the place of patients in the health care system as well as in the therapeutic relationship [15]. In the hospital, some patients receive information about their illness and a good understanding of their treatment. We have understood that informing the patient does not guarantee this latter a full knowledge of his or her state of health. According to the EUROPEP questionnaire, which is based on research by Gould et al., there are five dimensions to be considered inpatient satisfaction: the *doctor-patient relationship, medical care, information* and *support for the patient, continuity of care and the organization of consultations* [*16*]. The “Fédération Maison médicale santé” for its part, mentions two determinants of satisfaction: those related to the patient’s characteristics and those related to the health care received [16]. As the following patient states, the majority of patients are satisfied with the relationship they have with the doctors (Table.:4). *I am very confident and satisfied with my relationship with the doctors*. Patient of the surgery department.

If morality is a set of rules of conduct, social relations that society gives itself, which vary according to the culture, beliefs, living conditions and needs of the society, Frédéric Worms speaks of moral relations by calling them “relations between” as opposed to “relations to”. He contrasts them by speaking of the latter as an essentially spatial relationship between two objects and the relations between as characterized by the presence of affective behavior. Between doctor and patient, this affective behavior manifests itself everywhere, for example by offering a good welcome, by showing empathy, by informing the patient of his or her condition, to say nothing else. A recent study at HUEH showed that 45% of patients found doctors non-empathetic and the vast majority were poorly or not at all informed [17] and this more or less parallels the results of our study.

### Limitations and strengths

The research was conducted with available and willing patients without predetermining which patients would be best able to provide the best pieces of information. The content of some interviews did not include all the variables considered and some poor quality recordings were eliminated. The use of the qualitative approach and the respect of anonymity allowed patients to express themselves more freely. The number of participants and visits to several services made the data more abundant, more varied and thus more representative of HUEH.

### Conclusion and recommendation

This study shows that at HUEH the dominant relational type is paternalism by this tendency of physicians to take charge of everything without encouraging active patient participation. We also conclude that, while according to Worms the relationships are always a mixture of the relationship between and relationship to, the latter is predominant which makes the doctor-patient relationship at HUEH a non-moral one. Our study, being the first to have been carried out on this subject at the Hospital of the State University of Haiti, will allow a revision or an adjustment of the health policy by the Ministry of Public Health. It is also in itself a way of raising awareness among health professionals in the country, particularly for better care of patients by establishing a dynamic relationship involving the different components, doctor and patient, and this at all levels of care.

Thus we take the opportunity to recommend that practicing physicians be made aware of the importance of seeking the patient’s informed consent, the increase in the number of psychologists in the hospital, to train doctors who are also educators for patients, making them aware of their rights and the role they can play in their personal care, and focusing the training of health care professionals so that they see the patient and not just the disease.

## Data Availability

Data availability is not applicable for this article, as no database was used. The information was collected via audio recordings.

## Acknowledgments

Thanks to all students graduating from the Faculty of Medicine and Pharmacy of the State University of Haiti

## Ethics approval

The work was approved by the ethics committee of the Faculty of Medicine and Pharmacy of the State University of Haiti (LABMES: Laboratory of Ethical Medicine and Society).

